# Centrifugal filtration of serum for FTIR spectroscopy does not improve stratification of brain tumours

**DOI:** 10.1101/2022.12.14.22283465

**Authors:** Ashton G. Theakstone, Paul M. Brennan, Michael D. Jenkinson, Royston Goodacre, Matthew J. Baker

## Abstract

Discrimination of brain cancer *versus* non-cancer patients using serum-based attenuated total reflection Fourier transform infrared (ATR-FTIR) spectroscopy diagnostics was first developed by Hands *et al*. Cameron *et al*. then went on to stratifying between specific brain tumour types: glioblastoma multiforme (GBM) vs. primary cerebral lymphoma. Expanding on these studies, 30 GBM, 30 lymphoma and 30 non-cancer patients were selected to investigate the influence on test performance by focusing on specific molecular weight regions of the patient serum. Membrane filters with molecular weight cut offs of 100 kDa, 50 kDa, 30 kDa, 10 kDa and 3 kDa were purchased in order to remove the most abundant high molecular weight components. Three groups were classified using both partial least squares-discriminate analysis (PLS-DA) and random forest (RF) machine learning algorithms; GBM *versus* non-cancer, lymphoma *versus* non-cancer and GBM *versus* lymphoma. For all groups, once the serum was filtered the sensitivity, specificity and overall balanced accuracies decreased. This illustrates that the high molecular weight components are required for discrimination between cancer and non-cancer as well as between tumour types. From a clinical application point of view, this is preferable as less sample preparation is required.

## Introduction

Brain cancer diagnosis is challenging. The most common symptoms are non-specific (such as headaches) and are more likely to be associated with a non-tumour diagnosis (1-3). As many as two thirds of patients are diagnosed in the Emergency Department when their symptoms have deteriorated, with the majority of these patients having previously visited their primary care doctor multiple times (4). There is a need for a rapid, cost-effective and non-invasive tool for earlier diagnosis.

A vibrational spectroscopic technique, attenuated total reflection (ATR) Fourier transform infrared (FTIR) spectroscopy, has been applied to earlier detection and diagnosis of brain tumours (5, 6). Serum-based ATR-FTIR combined with machine learning algorithms can reliably predict which patients with symptoms of a possible brain tumour actually have a tumour on brain imaging. Hands *et al*. were the first to investigate the use of serum for ATR-FTIR spectroscopic analysis for brain tumour diagnosis, comparing brain tumour and asymptomatic non-tumour patients. Subsequent studies have included symptomatic non-tumour patients as well as investigating predictions of tumour grade and subtype (7-9). The earlier work used a traditional, time-consuming ATR-FTIR set-up with a fixed-point diamond internal reflection element (IRE). A newer, high-throughput approach uses silicon-based IRE (SIRE) sample slides. These SIREs are disposable and have multiple sampling points, which allows for high-throughput and batch processing (10, 11).

With this technique, brain tumours can be detected with a sensitivity of 88.7%, specificity of 94.7% and overall balanced accuracy of 91.7% (12). To further improve test performance, we investigated whether specific molecular weight regions of patient serum improved detection and stratification. Blood serum contains over 20,000 different proteins with a wide range of molecular weights, dominated by human serum albumin (HSA); 30-50 g/L is considered normal (13). Imbalances within protein concentrations in serum may relate to specific disease states and the low molecular weight fraction of serum may contain cancer-specific diagnostic information (14, 15).

Commercially available centrifugal filters can fractionate serum according to a molecular threshold, and so aid investigation of specific molecular weight fractions of serum. Traditionally, these filters are used to separate and remove rapidly the most abundant high molecular weight proteins from the less abundant low molecular weight molecules (*viz*. metabolites). One concern with these filters is the extra sample preparation required, the binding of small molecules to proteins which are then removed by filtrations. The reported potential for contamination from the filter membrane, has been resolved by Bonnier *et al*. who developed a centrifugal washing technique to remove any trace glycerine from the filter membranes (16, 17).

Here, we use the serum-based ATR-FTIR technique to investigate six (five fractions plus unfiltered whole serum) different molecular weight regions of serum for the stratification of brain cancer patients against non-cancer controls (Hands *et*.*al* previously reported sensitivity of 92.8% and specificity of 91.5% (7)).. We also explore the stratification between tumour types; GBM and primary cerebral lymphoma (Cameron *et al*. previously reported sensitivity of 90.1% and specificity of 86.3% (10)).

## Materials and Methods

Patient serum samples (*n*=90) were obtained from the Walton Centre NHS Trust (Liverpool, UK) and the Royal Preston Hospital (Preston, UK) with consent, under Ethics approval code (Walton Research Bank BTNW/WRTB 13_01/BTNW Application #1108). Included within the study were 30 glioblastoma (GBM) patients, 30 primary cerebral lymphoma patients and 30 asymptomatic control patients.

The patient serum was fractionated sequentially through five different size molecular weight filters (100 kDa, 50 kDa, 30 kDa, 10 kDa and 3 kDa) (Amicon Ultra-0.5 mL, Merck, Germany). The samples were centrifuged at 14,000 x*g* for 30 min to collect molecular weight fractions. The filtrate was collected and analysed so each portion represented the molecular weights less than the cut-off point (E.g., <100 kDa). This resulted in 6 serum samples per patient (including unfiltered whole serum), with a total of 540 samples.

Before the filters were used, they were centrifugally washed with 0.1M NaOH and MilliQ water through the following steps; 30 min with 0.1M NaOH at 14,000 x*g*, followed by 2 times 30 min with MilliQ water at 14,000 x*g*, and finally 2 min upside down at 1,000 x*g* to remove any remaining liquid. The washing was necessary to remove any residual glycerine coating on the ultrafiltration membranes as indicated by the manufacturer, to ensure no interferences within the sample spectra. Within the Supplementary Information there is example serum spectra illustrating both washed and unwashed filters, highlighting the need for the pre-analytical washing steps (Figure S1).

Patient serum, either whole or molecular weight fraction (3 μL), was deposited onto a SIRE optical sample slide (Dxcover Ltd, Glasgow, UK) and air dried before spectroscopic data collection. All serum spectra were collected on a Perkin Elmer Spectrum 2 FTIR spectrometer (Perkin Elmer, London, UK), utilising a Specac Quest ATR accessory unit with a specular reflectance puck (Specac Ltd., London, UK), allowing a Dxcover optical sample SIRE (Dxcover Ltd., Glasgow, UK) to be placed directly on top of the aperture. Each sample SIRE contains four wells where one remains blank as the background and the other three were used as sample repeats, with each three wells analysed three times. Nine spectra per patient were collected within the range of 4000–450 cm^−1^, at a resolution of 4 cm^−1^, with 1 cm^−1^ data spacing and 16 co-added scans; resulting in a total of 4,860 spectra acquired. The typical time for spectral collection was 15 min per patient sample slide (9 repeats and background).

The spectroscopic data analysis was completed using the R Statistical Computing Environment, MATLAB R2020a software with the PRFFECT toolbox (18) or a principal component analysis (PCA) code written in house. Data pre-processing was applied to reduce computational burden and improve classification algorithms. The techniques used match previous published work (12), (19). Exploratory analysis was completed using PCA followed by supervised machine learning methods including random forest (RF) and partial least squares-discriminant analysis (PLS-DA). The supervised techniques require splitting the data into training and test sets where the training set is used to identify biosignatures in a calibration phase and the model generated subsequently used for predictions to be made on the test set (10, 11, 20, 21). As there were no imbalances between the groups, no training set sampling adjustments were needed for classification analysis. The three groups were classified as the following: (i) GBM *versus* non-cancer, (ii) lymphoma *versus* non-cancer and (iii) GBM *versus* lymphoma.

Each classification completed using the PRFFECT toolbox had 51 reiterations to minimise standard error and to ensure a robust diagnostic model was used. The data were randomly split by patient ID at a 70/30 ratio between the training and test sets, keeping all patient spectral repeats together. The 51 reiterations shuffled the 70/30 split each time so that every patient within the whole dataset was predicted at least once.

## Results

Figure 1 displays the spectral differences between the same GBM patient sample in unfiltered serum compared to each of the five molecular weight cut-off regions. Each patient serum was separated through a 100 kDa filter first, followed by 50 kDa, then 30 kDa, followed by 10 kDa and finally 3 kDa, where the filtrate (region that has passed through the filter) was analysed. This process resulted in the molecular weight regions of <100 kDa, <50 kDa, <30 kDa, <10 kDa and <3 kDa. From Figure 1, it is clear that there is a large difference within the serum spectra once the higher molecular weight components (>100 kDa) were removed. This is significant in the higher wavenumber region between 3700 cm^-1^ to 2700 cm^-1^. However, more importantly there are numerous differences between 1800 cm^-1^ and 1000 cm^-1^ (Figure 1 inset), which was therefore determined as the region of interest for further analyses. The Amide I and II bands are reduced after filtration, which is perhaps unsurprising as HSA and other serum-based proteins have been removed (note HSA is ∼50% of the protein content of human blood). Given this observation, as to be expected, there are no visual differences observed between the three groups of patients; GBM, lymphoma and non-cancer (Figure S2 and S3), however they all followed the same trend as Figure 1 once separated into specific molecular weight fractions.

**Figure 1:**
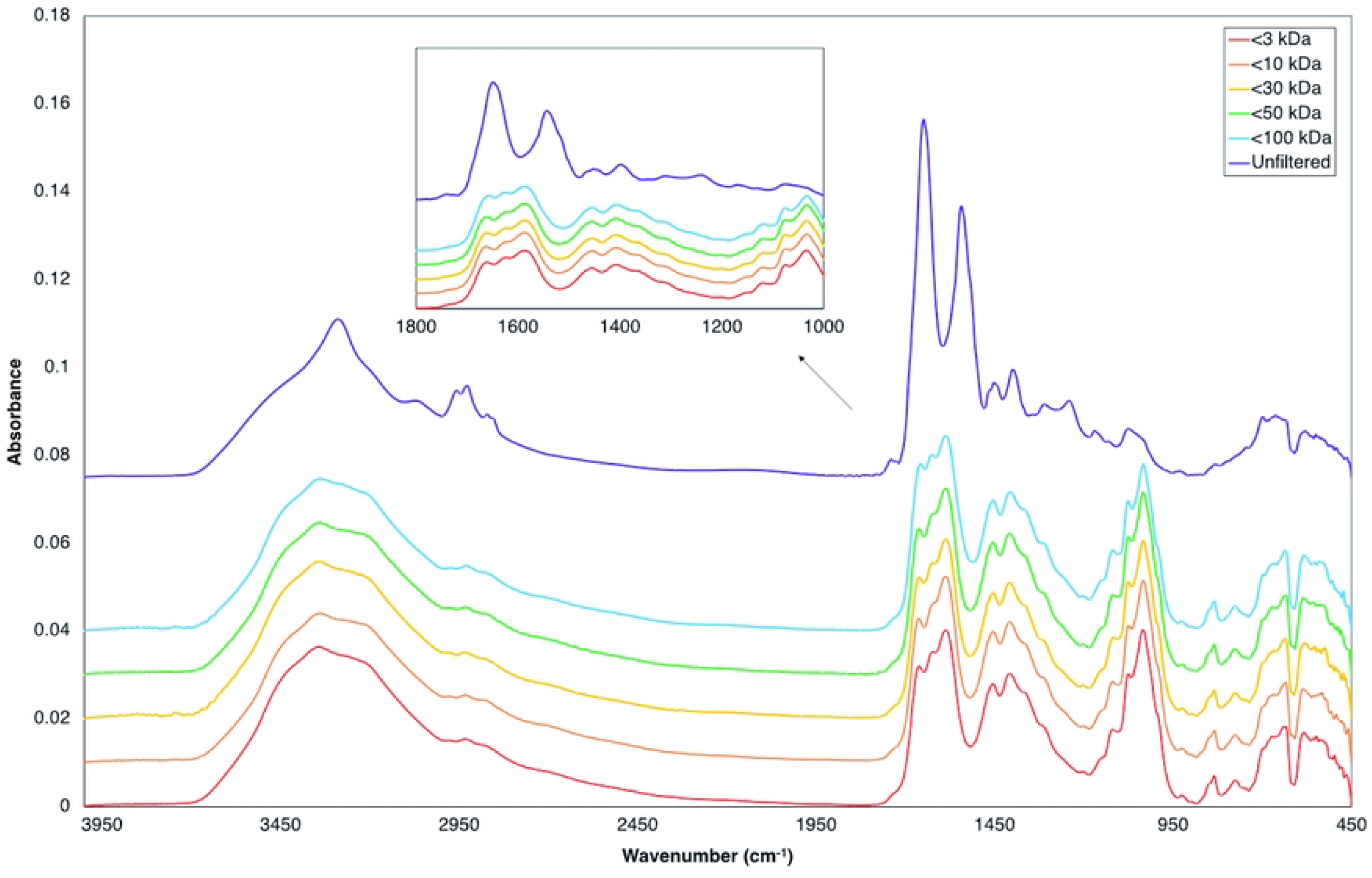
Example of patient serum spectra including unfiltered whole serum and each molecular weight region. Average of the 30 GBM patients shown here. The inset is the wavenumber region between 1800 cm^-1^ and 1000 cm^-1^, which was used for all chemometrics and machine learning analyses. Spectra is offset for clearer visualisation.

The patient spectral data were subjected to both exploratory PCA and supervised classification models (RF and PLS-DA) for all three groups: (i) GBM *versus* non-cancer, (ii) lymphoma *versus* non-cancer and (iii) GBM *versus* lymphoma. Figure 2 illustrates the PCA scores results for the three groups with unfiltered serum and contains a slight separation between the classes along the second principal component (PC2). Figure 3 shows the PCA outcomes for the three groups in the molecular weight region <100 kDa, where there is no clear separation between in the classes in all groups. The PCA scores plots for <50 kDa, <30 kDa, <10 kDa and <3 kDa are contained with the supplementary information (Figures S4-S7) and display similar results to that of the <100 kDa region, with no separation between the classes (GBM *versus* non-cancer, lymphoma *versus* non-cancer and GBM *versus* lymphoma).

**Figure 2:**
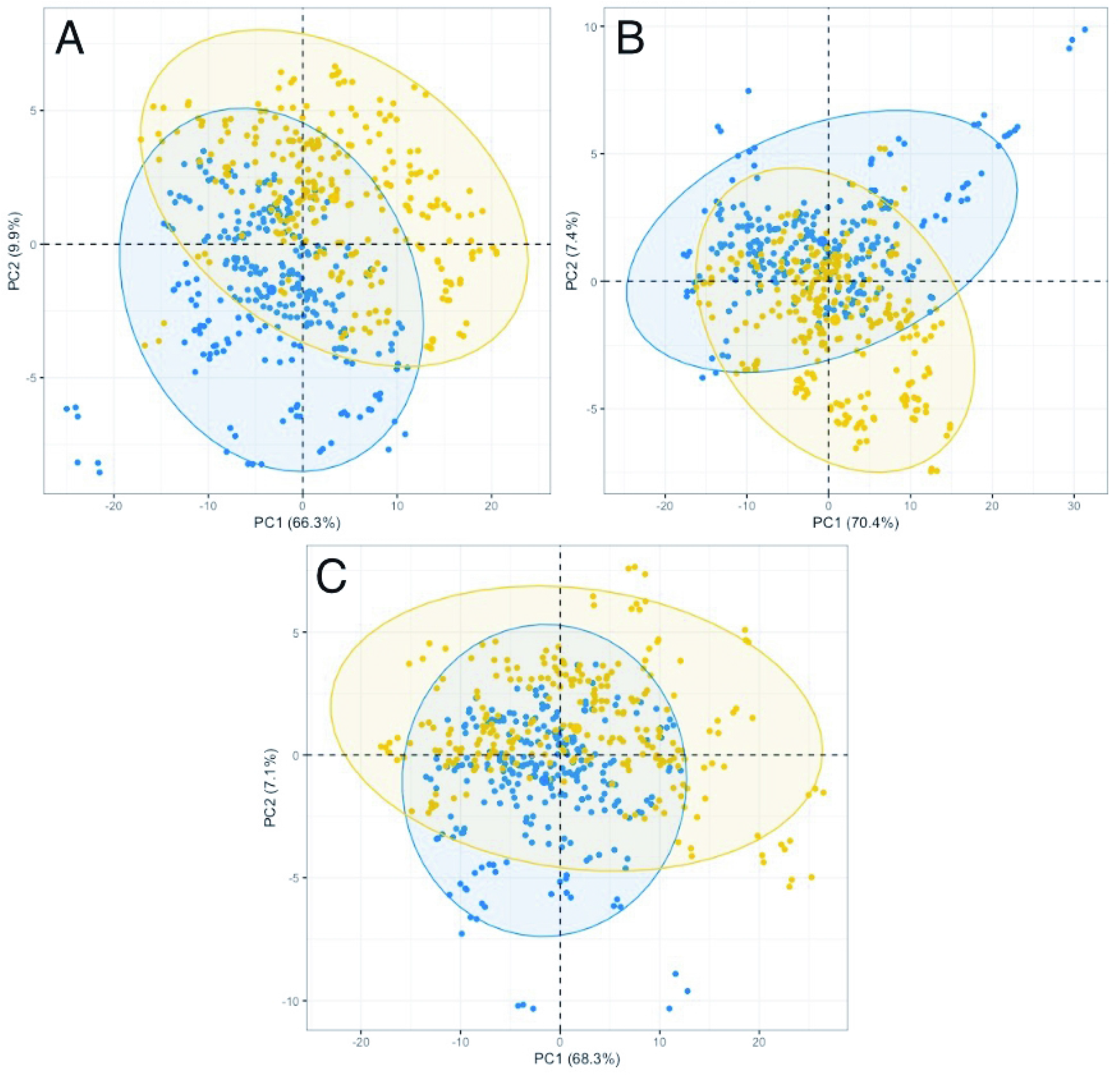
Principal component analysis scores plots for the unfiltered whole serum of the first and second dimensions. The three figures represent (A) GBM in blue and non-cancer in yellow, (B) lymphoma in blue and non-cancer in yellow and (C) GBM in blue and lymphoma in yellow. The eclipses in each class represent a 95% confidence interval. Values in parentheses within the axes legends are the total explained variance (TEV) for each principal component (PC).

**Figure 3:**
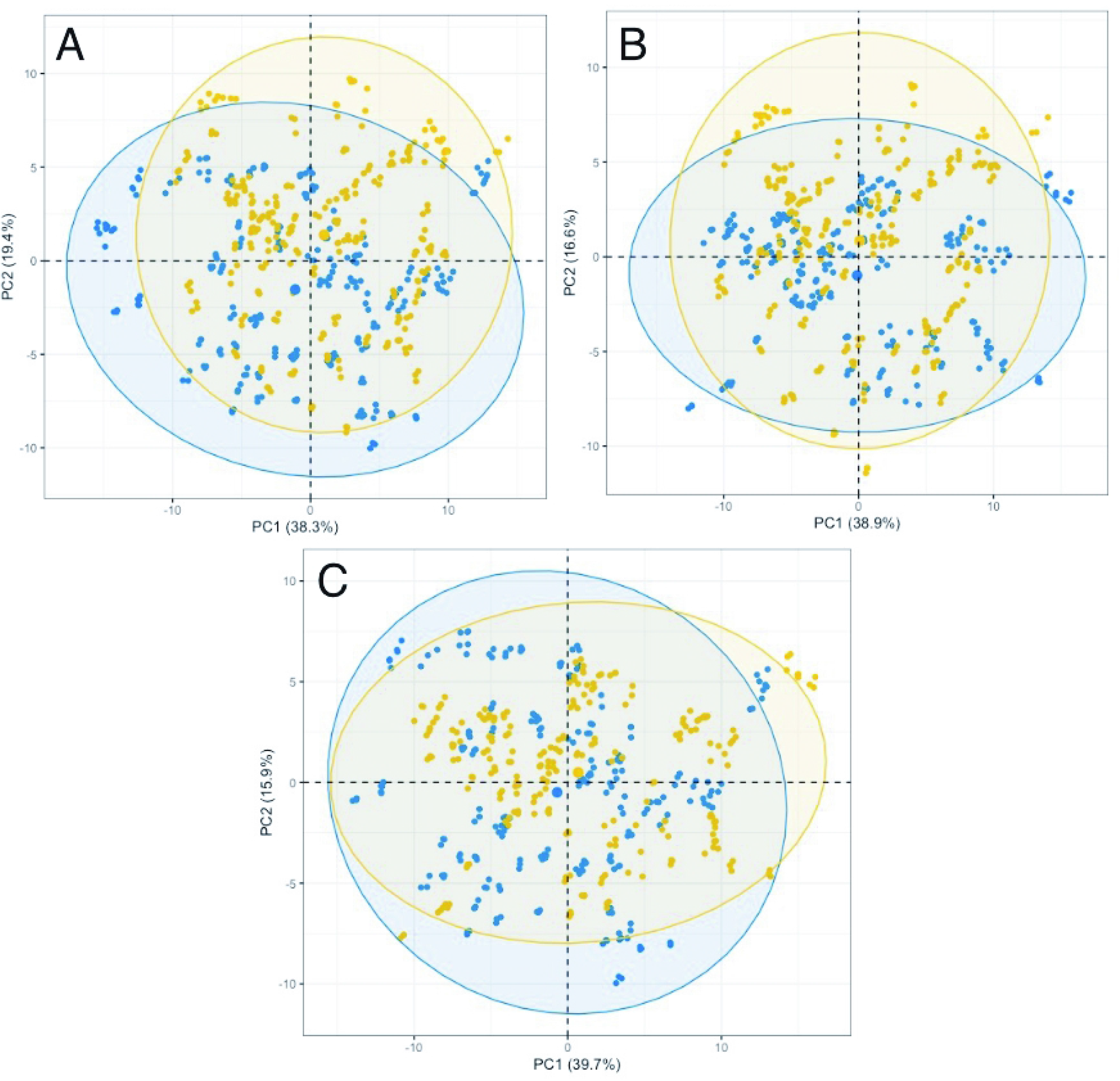
PCA scores plots for the filtered serum (<100 kDa) of the first and second dimensions. The three figures represent (A) GBM in blue and non-cancer in yellow, (B) Lymphoma in blue and non-cancer in yellow and (C) GBM in blue and lymphoma in yellow. The eclipses in each class represent a 95% confidence interval. Values in parentheses are the TEV for each PC.

Following this initial exploratory PCA, each group was analysed using the supervised learning algorithms of RF and PLS-DA. Tables 1-3 contain the sensitivity, specificity and balanced accuracy (defined as the averaged sensitivity and specificity) for each serum fraction. The PLS-DA model results are included within these tables (Tables 1-3), while the RF are provided within the supplementary information (Tables S1-S3).

**Table 1:** Sensitivity, specificity and balanced accuracies for the PLS-DA model classification of GBM *versus* non-cancer patients. Mean, standard deviation (SD) and 95% confidence intervals (CIs) are provided.

|  | Sensitivity (%) |  |  | Specificity (%) |  |  | Balanced accuracy (%) |  |  |
| --- | --- | --- | --- | --- | --- | --- | --- | --- | --- |
|  | Mean | SD | 95% CI | Mean | SD | 95% CI | Mean | SD | 95% CI |
| Unfiltered | 87.4 | 11.8 | $\pm 3.2$<br>84.2-90.6 | 92.4 | 8.2 | $\pm 2.3$<br>90.1-94.7 | 89.9 | 6.4 | $\pm 1.8$<br>88.1-91.7 |
| <100 kDa | 85.5 | 10.7 | $\pm 2.9$<br>82.6-88.4 | 92.4 | 8.2 | $\pm 2.3$<br>90.1-94.7 | 89.0 | 6.0 | $\pm 1.6$<br>87.4-90.6 |
| <50 kDa | 84.3 | 11.5 | $\pm 3.2$<br>81.1-87.5 | 88.9 | 10.4 | $\pm 2.9$<br>86.0-91.8 | 86.6 | 6.3 | $\pm 1.7$<br>84.9-88.3 |
| <30 kDa | 84.0 | 13.9 | $\pm 3.8$<br>80.2-87.8 | 85.6 | 12.4 | $\pm 3.4$<br>82.2-89.0 | 84.8 | 8.7 | $\pm 2.4$<br>82.4-87.2 |
| <10 kDa | 79.7 | 12.7 | $\pm 3.5$<br>76.2-83.2 | 85.2 | 13.5 | $\pm 3.7$<br>81.5-88.9 | 82.4 | 7.4 | $\pm 2.0$<br>80.4-84.4 |
| <3 kDa | 86.8 | 13.7 | $\pm 3.8$<br>83.0-90.1 | 87.4 | 9.7 | $\pm 2.7$<br>84.7-90.1 | 87.1 | 8.3 | $\pm 2.3$<br>84.8-89.4 |

**Table 2:** Sensitivity, specificity and balanced accuracies for the PLS-DA model classification of lymphoma *versus* non-cancer patients. Mean, standard deviation (SD) and 95% confidence intervals (CIs) are provided.

|  | Sensitivity (%) |  |  | Specificity (%) |  |  | Balanced accuracy (%) |  |  |
| --- | --- | --- | --- | --- | --- | --- | --- | --- | --- |
|  | Mean | SD | 95% CI | Mean | SD | 95% CI | Mean | SD | 95% CI |
| Unfiltered | 85.3 | 13.6 | $\pm 3.7$<br>81.6-89.0 | 85.8 | 13.3 | $\pm 3.7$<br>82.1-89.5 | 85.6 | 8.7 | $\pm 2.4$<br>83.2-88.0 |
| <100 kDa | 66.1 | 18.5 | $\pm 5.1$<br>61.0-71.2 | 78.9 | 14.9 | $\pm 4.1$<br>74.8-83.0 | 72.5 | 11.5 | $\pm 3.2$<br>69.3-75.7 |
| <50 kDa | 66.9 | 19.7 | $\pm 5.4$<br>61.5-72.3 | 83.7 | 12.8 | $\pm 3.5$<br>80.2-87.2 | 75.3 | 10.9 | $\pm 3.0$<br>72.3-78.3 |
| <30 kDa | 74.2 | 17.4 | $\pm 4.7$<br>69.4-79.0 | 80.6 | 13.8 | $\pm 3.8$<br>76.8-84.4 | 77.4 | 10.4 | $\pm 2.9$<br>74.5-80.3 |
| <10 kDa | 70.6 | 16.4 | $\pm 4.5$<br>66.1-75.1 | 81.5 | 12.1 | $\pm 3.3$<br>78.2-84.8 | 76.0 | 10.8 | $\pm 3.0$<br>76.0-79.0 |
| <3 kDa | 65.7 | 15.2 | $\pm 4.2$<br>61.5-69.9 | 82.8 | 13.6 | $\pm 3.7$<br>79.1-86.5 | 74.2 | 9.6 | $\pm 2.6$<br>71.6-76.8 |

**Table 3:**
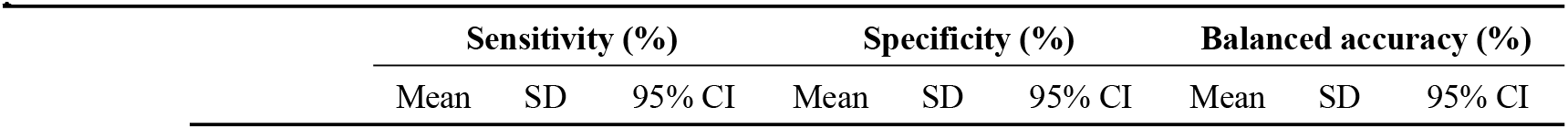
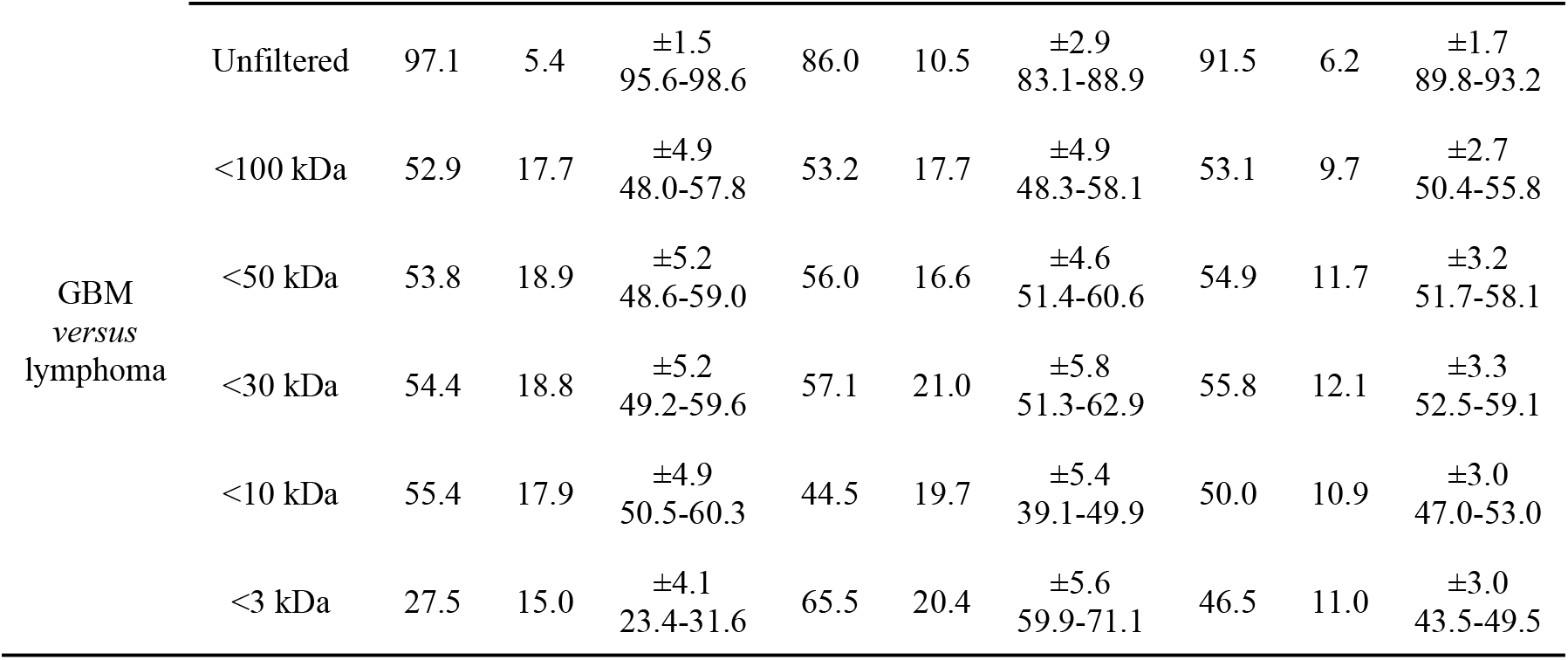
Sensitivity, specificity and balanced accuracies for the PLS-DA model classification of GBM *versus* Lymphoma patients. Mean, standard deviation (SD) and 95% confidence intervals (CIs) are provided.

| Sensitivity (%) |  |  | Specificity (%) |  |  | Balanced accuracy (%) |  |  |
| --- | --- | --- | --- | --- | --- | --- | --- | --- |
| Mean | SD | 95% CI | Mean | SD | 95% CI | Mean | SD | 95% CI |

|  |  |  |  |  |  |  |  |  |  |  |
| --- | --- | --- | --- | --- | --- | --- | --- | --- | --- | --- |
| GBM<br>*versus*<br>lymphoma | Unfiltered | 97.1 | 5.4 | $\pm 1.5$<br>95.6-98.6 | 86.0 | 10.5 | $\pm 2.9$<br>83.1-88.9 | 91.5 | 6.2 | $\pm 1.7$<br>89.8-93.2 |
| | <100 kDa | 52.9 | 17.7 | $\pm 4.9$<br>48.0-57.8 | 53.2 | 17.7 | $\pm 4.9$<br>48.3-58.1 | 53.1 | 9.7 | $\pm 2.7$<br>50.4-55.8 |
| | <50 kDa | 53.8 | 18.9 | $\pm 5.2$<br>48.6-59.0 | 56.0 | 16.6 | $\pm 4.6$<br>51.4-60.6 | 54.9 | 11.7 | $\pm 3.2$<br>51.7-58.1 |
| | <30 kDa | 54.4 | 18.8 | $\pm 5.2$<br>49.2-59.6 | 57.1 | 21.0 | $\pm 5.8$<br>51.3-62.9 | 55.8 | 12.1 | $\pm 3.3$<br>52.5-59.1 |
| | <10 kDa | 55.4 | 17.9 | $\pm 4.9$<br>50.5-60.3 | 44.5 | 19.7 | $\pm 5.4$<br>39.1-49.9 | 50.0 | 10.9 | $\pm 3.0$<br>47.0-53.0 |
| | <3 kDa | 27.5 | 15.0 | $\pm 4.1$<br>23.4-31.6 | 65.5 | 20.4 | $\pm 5.6$<br>59.9-71.1 | 46.5 | 11.0 | $\pm 3.0$<br>43.5-49.5 |

For the GBM *versus* non-cancer there was a slight decrease in the sensitivity, specificity and balanced accuracies once the serum was filtered. However, all classification models had an overall balanced accuracy greater then 82%, suggesting that even the individual molecular weight regions of serum can predict GBM from non-cancer patients. Lymphoma *versus* non-cancer had a larger decrease in sensitivity, specificity and balanced accuracies once filtered with overall balanced accuracies ranging between 72% and 78%. When investigating between the two cancer types, GBM *versus* lymphoma, there was a significant decrease in the sensitivity, specificity and balanced accuracies. The overall balanced accuracy decreased from 91.5% to a range between 46% and 56%, suggesting the serum fractions are unreliable in being able to stratify between cancer types as there is no distinction between GBM and lymphoma.

The RF classifications for all three groups gave very similar responses to the PLS-DA. For GBM *versus* non-cancer there was more of a decrease in sensitivity, specificity and balanced accuracies once the serum was filtered. The same can be said with Lymphoma *versus* non-cancer and once again, there was no ability to stratify between the cancer types using RF model algorithms. The percentages for each groups sensitivity, specificity and balanced accuracies are displayed in the supplementary information (Tables S1-S3).

From these PLS-DA classification models the loadings plots were investigated in order to identify which wavenumber regions were important for the discriminations between the cohorts. Figures 4 and 5 display the PLS-DA loadings plot for each group with the unfiltered serum (Figure 4) and the first fraction of filtered serum (<100 kDa) (Figure 5). Both the first and second PLS components are shown within the figures as the majority of the spectral variance between the cohorts will be present within these two latent variables. The loadings plots for the other serum fractions (<50 kDa, <30 kDa, <10 kDa and <3 kDa) are included within the supplementary information (Figures S8-S11).

**Figure 4:**
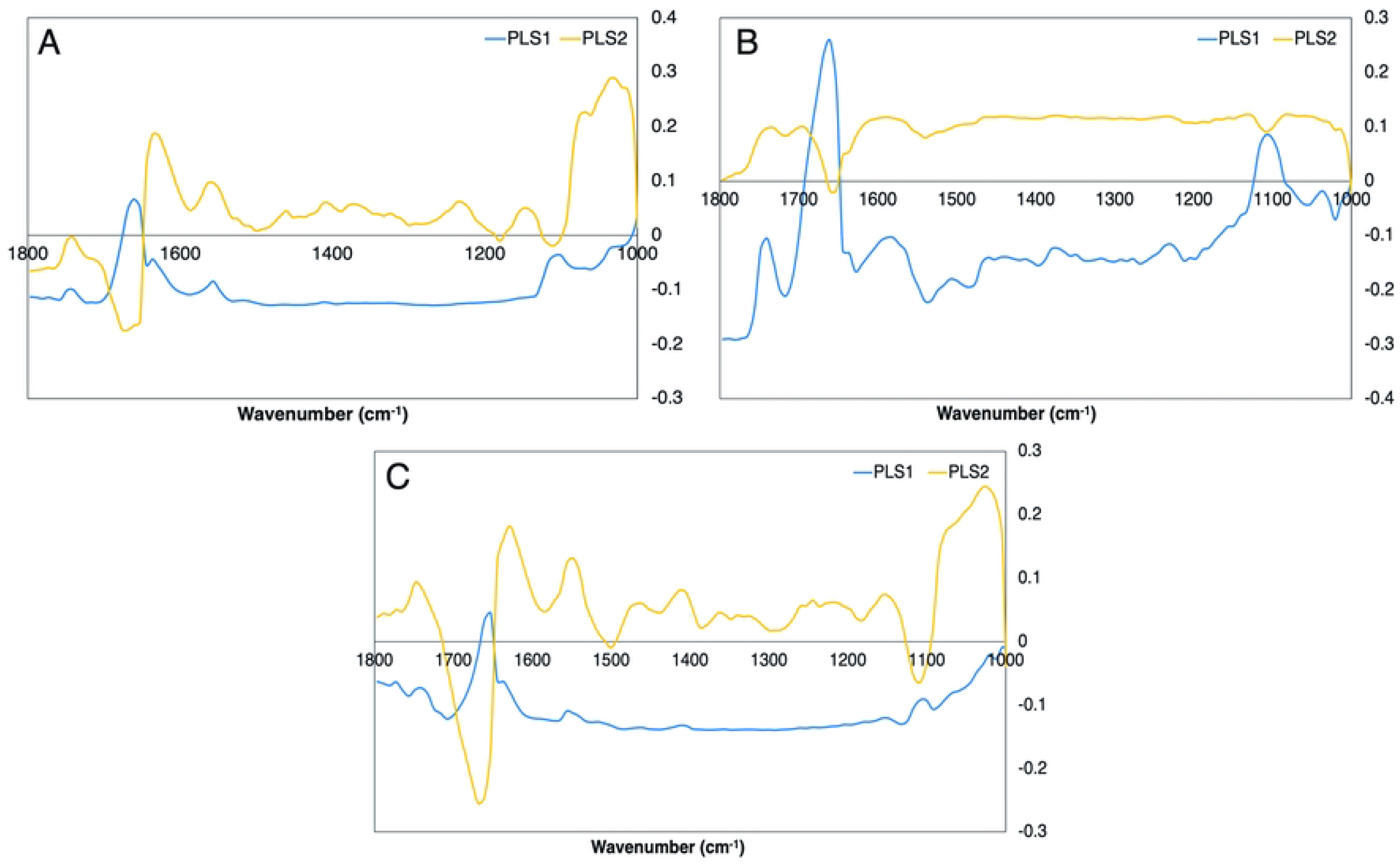
PLS loadings plots for the 1^st^ and 2^nd^ latent variables (LVs) for the unfiltered whole serum. (A) GBM *versus* non-cancer, (B) Lymphoma *versus* non-cancer and (C) GBM *versus* lymphoma.

**Figure 5:**
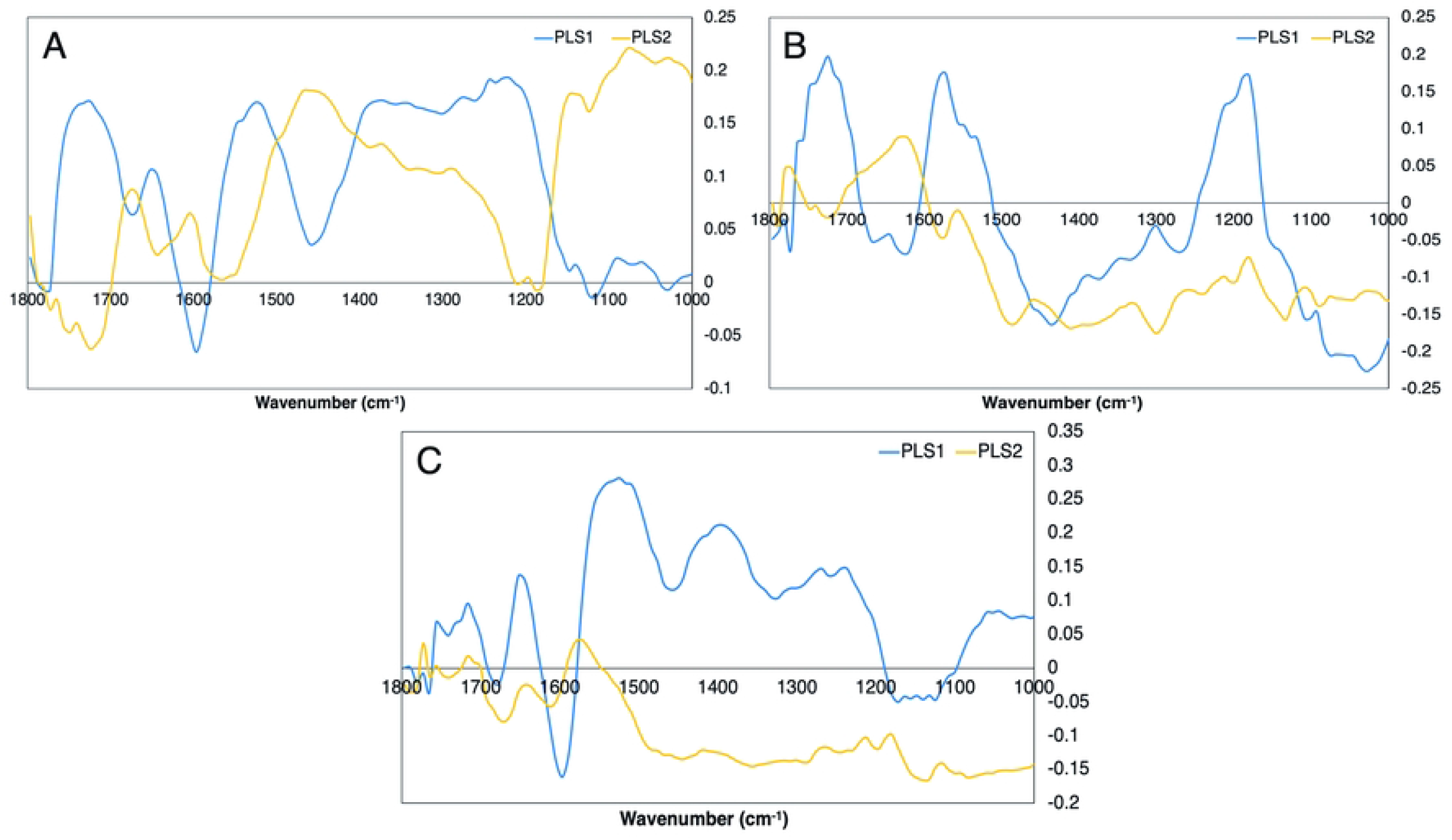
PLS loadings plot for the 1^st^ and 2^nd^ LVs for the filtered serum (<100 kDa). (A) GBM *versus* non-cancer, (B) Lymphoma *versus* non-cancer and (C) GBM *versus* lymphoma.

For the unfiltered serum the loadings plot suggests that the discrimination between GBM and non-cancer (Figure 4.A) is dependent on the Amide I and Amide II proteins (region between 1700 cm^-1^ and 1500 cm^-1^) and the glycogen/carbohydrates (1100 cm^-1^ – 1000 cm^-1^). Lymphoma *versus* non-cancer (Figure 4.B) has similar reliance on the Amide I and Amide II bands, however there is less importance in the glycogen/carbohydrates. The peak at ∼1740 cm^-1^ suggests that the discrimination between lymphoma and non-cancer is also determined by the lipid components within the serum. Discriminating between cancer types, GBM *versus* lymphoma (Figure 4.C), there is importance within the Amide I and Amide II region, and the glycogen/carbohydrates region. The peaks identified as important for each group is displayed in Table 4.

**Table 4:**
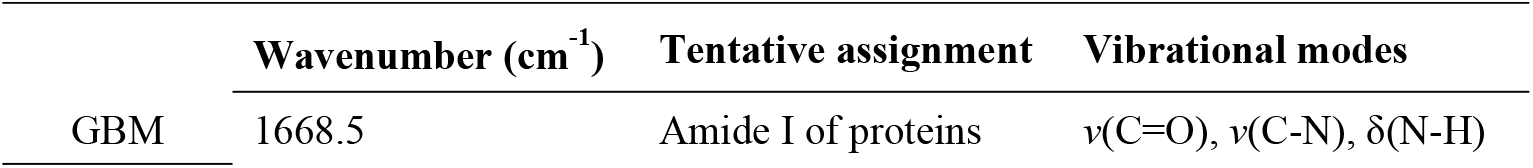
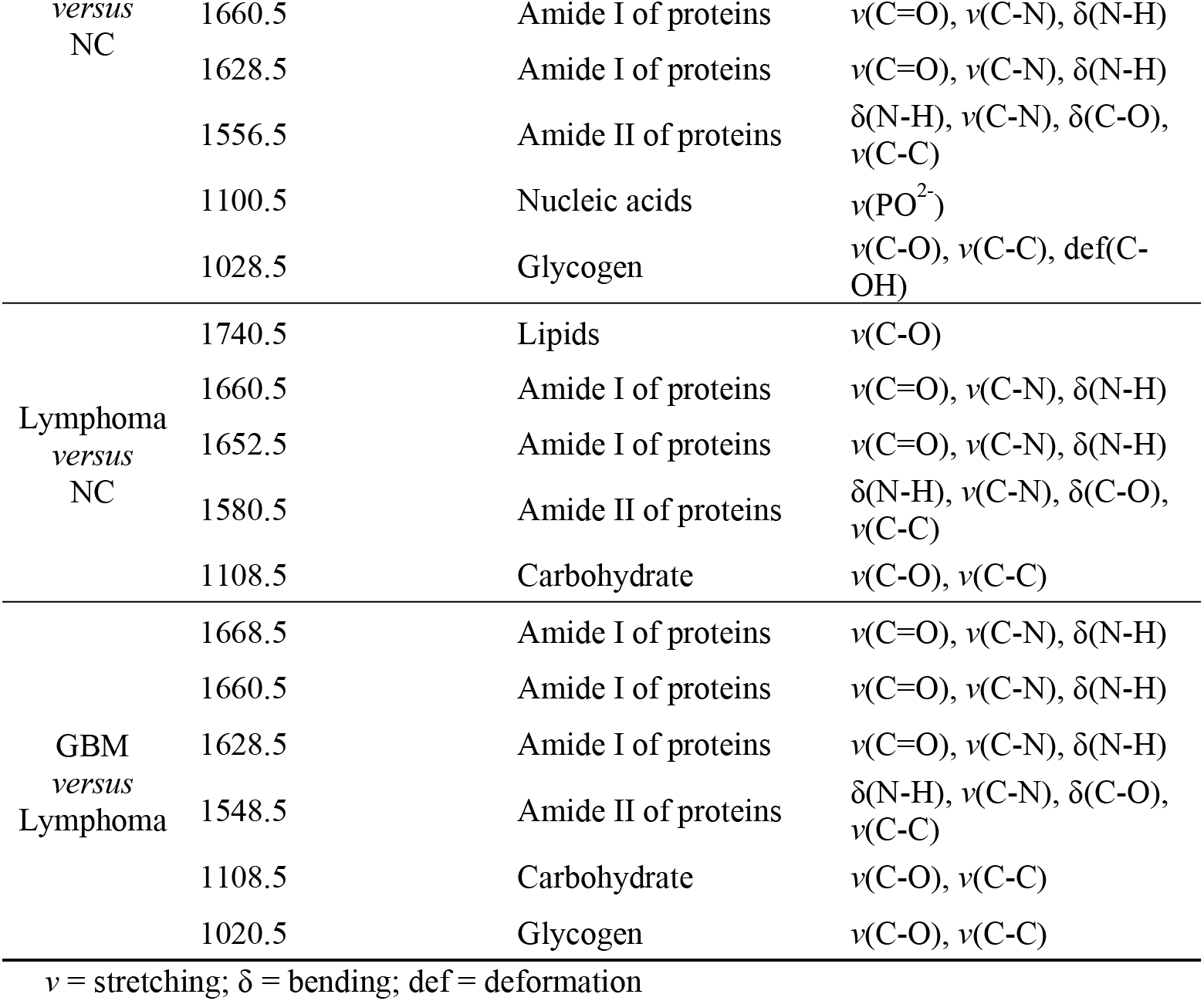
Top wavenumbers for each group in unfiltered serum classifications. Tentative biochemical assignments and their corresponding vibrational modes are included (22).

Once the serum was filtered the loadings plot significantly changed with what wavenumber regions were deemed important for the discrimination between patient cohorts (Figure 5). The percentage of variance from each LV will decrease with the accuracy of the model, therefore it is unreasonable to directly compare the important peaks from filtered and unfiltered serum when the whole serum will have a greater percentage of importance. It is interesting to note that more regions are deemed important within the filtered serum however the percentage of variance represented in each LV is minimal compared to the whole serum.

## Discussion

From the initial visual observations there was a significant difference between the spectral profiles of whole unfiltered serum and the different molecular weight fractions. This is to be expected as the initial filtration step will remove components greater than 100 kDa, including human serum albumin which comprises 50% of the protein complement of sera (30-50 g/L), and antibodies such as immunoglobulin G (IgG). IgG is one of the main components (7-16 g/L) within serum (23, 24). The removal of serum albumin and other components within the first filtration step has a significant impact on the overall serum spectral profile. Between the 3 patient groups, GBM, lymphoma and non-cancer, visually there were no spectral differences and they all follow the same spectral trend once centrifugally filtered (Figures 1, S2 and S3). There were visually few changes between the molecular weight fractions as most significant changes occured within the first filtration step.

Within the exploratory principal component analysis (PCA) there was slight separation between the groups along the second principal component for the unfiltered whole serum. By contrast, once filtered there was no separation between the groups of patients, demonstrated throughout all molecular weight regions. The clear distinction between groups within the unfiltered serum suggests that the higher molecular weight (>100 kDa) components within the serum play an important role for the discrimination between cancer and non-cancer or between cancer types.

These observations can be confirmed through the supervised classification analysis where each group of patients was stratified using both PLS-DA and RF machine learning algorithms. For the unfiltered serum the classifications between the two patient groups (GBM *versus* non-cancer, lymphoma *versus* non-cancer or GBM *versus* lymphoma) all gave overall balanced accuracies above 85%. When focusing on GBM *versus* non-cancer the sensitivities, specificities and balanced accuracies of the models remained around or greater then 80%; however, none of the molecular weight filtrates gave percentages as high as the unfiltered whole serum. Lymphoma *versus* non-cancer had a more noticeable decrease in sensitivity, specificity and balanced accuracies once the patient serum was filtered. These remained at 65% and greater, however, as with the GBM *versus* non-cancer cohort the unfiltered serum outperformed each filtrate. Between the two cancer types, GBM versus lymphoma, there was a significant decrease in stratification ability once the serum was filtered. The overall balanced accuracies of 50% suggest that there are no discriminatory features within the serum to identify between a GBM or lymphoma brain cancer patient once the components above 100 kDa were removed.

For all classifications the unfiltered whole serum performed the greatest which suggests that the higher molecular weight components are needed for discriminatory ability between these binary cohorts. From a clinical application point of view, this is preferable as the extra pre-analytical steps to include the filtration is more time consuming and harder to translate into a clinic ready test.

## Data Availability

All relevant data are within the manuscript and its Supporting Information files.

## Acknowledgements

The authors would like to thank both the Walton Centre NHS Foundation Trust and the Lancashire Teaching Hospitals NHS Trust for the collaboration and access to patient samples. This research was funded by Cancer Research UK (Grant number A28345). The patients were collected from either the Walton Centre or the Royal Preston Hospital are under ethics approval code (Walton Research Bank BTNW/WRTB 13_01/BTNW Application #1108).

